# Associations between Social Contact, Sleep and Dietary Patterns Among Children: A Cross-Sectional Study

**DOI:** 10.1101/2023.11.23.23298946

**Authors:** Christophe Mühlematter, Matthieu Beaugrand, Andjela Markovic, Salome Kurth

## Abstract

Social isolation in adults can be associated with altered sleep and eating behavior. This study aimed to investigate the interactions between the extent of social contact, eating behavior and sleep in infants and preschool children. In an observational study, 439 caregivers of 562 children aged 0-6 years provided information on sleep (i.e., duration, latency, bedtimes and nighttime awakenings), eating behaviors (i.e., meal size, consumption of sweet snacks, salty snacks, fruits and vegetables) and social contact (i.e., quarantine status, household size, social activities) during the Covid-19 pandemic (April 2020). In infants (0-3 years), the change in meal size and consumption of snacks, fruits, and vegetables did not significantly relate to the extent of social contact. For preschool children (3-6 years), a trend was observed, suggesting that quarantine status was associated with increased meal size. Changes in sleep duration, sleep latency, bedtimes and nighttime awakenings from before to during the pandemic were not significantly linked to the 3 variables quantifying social contact in both age groups. The study highlights that, contrary to expectations, the extent of social contact has negligible associations with infants’ and preschool children’s sleep and eating behaviors. These findings indicate that other factors beyond social isolation play a role in shaping children’s eating habits and sleep patterns.

## 1. Introduction

Social relationships are fundamental to well-being, mental and physiological health in adults (Cacioppo et al. 2002; Holt-Lunstad et al. 2015) and adolescents (Qualter et al. 2013). Reported associations between social relationships and physical health mainly focus on two constructs: social support and social integration. Social support represents the psychological and material resources provided by the social network to help cope with stress, while social integration represents the participation in social relationships (Cohen 2004). More generally, frequency and duration of social interactions have been linked with physical and mental health through changes in health-related behaviors, such as diet, exercise, or smoking (Berkman et al. 2000).

Social isolation in adults generates higher risk for depressive symptomology (Cacioppo, Hawkley, and Thisted 2010), mortality (Holt-Lunstad et al. 2015) and reduced sleep quality (Azizi-Zeinalhajlou et al. 2022; Cacioppo et al. 2002). One relevant factor intertwined with these observations may lie in altered eating behavior, based on indications that social isolation can be associated with eating disorders and binge eating (Mason et al. 2016; Wright and Pritchard 2009). The effect of social isolation on sleep and eating behavior has been experimentally examined in Drosophila, demonstrating that separating adult flies from the group led to increased food consumption and shorter sleep (Li et al. 2021).

The COVID-19 pandemic created a unique quasi-experimental framework to investigate the effects of enforced confinement on a global scale, downsizing direct social contact within populations. In this framework, social isolation significantly impacted mental health of children and parents (Ehrler et al. 2022). In addition, children and adolescents consumed more snacks and processed foods during the pandemic, leading to weight gain (González-Monroy et al. 2021; Pujia et al. 2021). Children and adolescents also experienced shortening of sleep (Markovic et al. 2021; Teixeira et al. 2021). Yet controversial results remain, for example as part of the COVID-19 pandemic, an increase in fruit and vegetable consumption was reported in Italian children ages 5 to 14 years (Pujia et al. 2021). The pandemic-induced reduction of social contacts could explain the changes in eating and sleeping behavior towards more “unhealthy” patterns, which aligns with findings from observational studies on social isolation. Indeed, the contrasting findings might be explained through differences in effective social isolation determined by the household structure, family size, extent of isolation orders by country, or geography (urban vs. rural regions). Thus, quantifying individual levels of social contact might contribute to a more precise understanding of the interactions among social contact, eating and sleep behaviors in children. Moreover, these interactions have not yet been examined in preschool-age children, who might respond differently to changes in social contacts due to their reliance on interactions with caregivers rather school-age children.

In this investigation, we thus tested the hypothesis that infants and preschool children who experienced increased social isolation (i.e., being in quarantine, engaging in activities alone rather than together with other family members and small household size) have worsened sleep quality (shorter sleep duration, increased night awakenings, longer sleep latency), and more “unhealthy” eating patterns (increased meal size, decreased consumption of fruits and vegetables, and increased consumption of sweet and salty snacks).

## 2. Materials and Methods

### 2.1. Study Design

During the start of the COVID-19 pandemic from April to July 2020, caregivers of children below 6 years were invited to participate in a study about children’s sleeping behavior. Recruitment was realized through large-scale distribution of digital flyers and a video invitation, distributed on social media platforms, childcare institutions, medical practices, and personal channels. 439 primary caregivers (423 females, 96.35%, age 38.28 ± 4.74 years, mean ± SD) completed an online survey (SoSci Survey; Leiner, 2021) with questions about their children’s demographic variables, sleeping behavior, eating behavior and social isolation. The survey was compiled in English, and then translated to Italian, Spanish, French, and German by the authors, which was then checked by at least two native speakers. For families with multiple children, questions were looped to assess data for each child individually. We collected a total dataset of 309 (146 females, 47.25%) infants and toddlers (between 0 and 35 months old), and 253 (127 females, 50.2%) preschool children (age 36 to 72 months). The institutional ethics board of the University of Fribourg, Switzerland, approved the study, and parents gave informed consent before completing the survey.

### 2.1. Sleep Behavior change

According to age, children’s sleep was either assessed with the Brief Infant Sleep Questionnaire (BISQ; 0-35 mo) (Sadeh, 2004) or with the Children’s Sleep Habits Questionnaire (CSHQ; 36-72 mo) (Owens et al., 2000). Caregivers reported on the child’s sleep behavior before (retrospectively) and during the confinement. The change in four aspects of sleep behavior was analyzed by means of a difference during-before the lockdown: sleep duration, sleep latency, bedtimes and nighttime awakenings (as in Markovic et al., 2021). For the infant group (0-36 months), parents reported infants’ sleep duration for nighttime and daytime sleep, which were summed for the total sleep time (in minutes), number of nighttime awakenings, sleep latency (in minutes), and bedtimes (in minutes). In the preschool group (36-72 months), sleep duration was quantified from the item “*My child sleeps about the same duration each 24-h-day (nighttime sleep and naps combined)*”, sleep latency from the item “*My child falls asleep within 20 min after going to bed*”, nighttime awakenings from “*How often does your child wake up during the night*” and bedtime from “*My child goes to bed at the same time at night*”. Each item for preschool children was captured with ratings from 1 to 5, such that 1 referred to never (being the case, *i*.*e*., at 0 days/week), 2 to rarely (1 day/week), 3 to sometimes (2-4 days/week), 4 to usually (5-6 days/week) and 5 to always (7 days/week) being the case.

### 2.2. Social contact

With three variables we captured the individual extent of social contact during the confinement: First, quarantine status indicating whether the caregiver was in quarantine at the time of survey completion. Second, household size indicating the number of people living in the household. And third, social activities where parents reported the duration their child engaged in several activities (e.g., watching television or playing), and whether the child was doing these activities alone or with someone else (parents, siblings or friends, for details see Appendix). The percentage of activity spent engaged with the parents compared to alone was computed. For example, a child who spent half of the time engaging in activities with someone else would be attributed a 50% participation rate in social activities.

### 2.3. Eating behavior change

We assessed the change in meal size from before the confinement to during the confinement with a 5-point Likert scale capturing whether the child’s overall meal size was “much smaller”, “a bit smaller”, “same as before”, “a bit larger” or “much larger”. Moreover, parents rated the child’s change in the consumption of salty and sweet snacks (between main meals), and the consumption of fruits and vegetables by means of “much less”, “a bit less”, “the same amount”, “a bit more” or “much more”.

### 2.4. Statistical Analysis

Statistical analyses were performed in R version 4.0.5 with packages dplyr (Wickham et al. 2021), and figures were created by using ggplot2 (Wickham 2016). Due to a small number of responses for meal-sizes “much smaller” (n = 3 for the infant group, n = 4 for preschool children) and “much larger” (n = 6 for infants, n = 4 for preschool children), the scales were consolidated into three response groups “decreased”, “no change”, “increased”. Accordingly, the consumption of salty and sweet snacks, fruits and vegetables were composed into a 3-point scale (“decreased”, “no change”, “increased”). In each model employed, the number of participants was adjusted for the exclusion of missing data. To examine the relationship between the extent of social contact and sleep behavior, we separately computed generalized linear models for the two age groups, with sleep duration, sleep latency, bedtimes and nighttime awakenings as the outcome and the extent of social contact (quarantine status, household size, social activities) as the predictor. For both age groups, the association between the extent of social contact and eating was tested with change in meal size, snacks, fruits and vegetables as outcome and social contact as predictor. Age and sex were included as control factors, and parental stress was included as a covariate in alignment with previous work demonstrating an association with children’s sleep (Markovic et al. 2021). Change in parental stress was captured with ratings from 1 to 5 with the question “*Did your behavior change across the time that passed since the lockdown, such that your level of stress: 1 decreased a lot, 2 decreased a little, 3 did not change, 4 increased a little, or 5 increased a lot*.” Alpha level was set to p < 0.05 and p-values were corrected for multiple testing using the false discovery rate method (Benjamini and Hochberg 1995).

## 3. Results

### 3.1. Study population

Demographic variables revealed that household size and percentage of quarantined families were similar in the age groups (Table 1). Percentage of shared activities was by 10.62% larger in the infant group compared to the preschool children. The control variable change in parental stress was comparable between groups.

**Table 1.** Demographic and control variables included in the analysis, indicated for the infant (n=309) and preschool-age group (n=253). Data are reported as mean and standard deviation.

| Demographic and control variables | Infants | Preschoolers |
| --- | --- | --- |
| Number of children | 309 | 253 |
| Number of girls | 146 (47.25%) | 127 (50.2%) |
| Age (years) | 1.58 ± 0.79 | 4.44 ± 0.8 |
| Change in parental stress | 0.47 ± 1.09 | 0.42 ± 1.17 |
| Social contact | Infants | Preschoolers |
| Household size (persons) | 3.72 ± 0.92 | 4.02 ± 0.89 |
| Quarantined families | 94 (30.42%) | 79 (31.23%) |
| Shared activities (percentage) | 76.09% ± 18.99 | 65.47% ± 19.19 |

### 3.2. Sleep behavior

Before the lockdown, sleep duration ranged from 570 to 1020 minutes for infants, during the lockdown it ranged between 510 and 990. Sleep latency before the lockdown was between 0 and 60 minutes, during the lockdown it was between 0 and 90 minutes. Before lockdown, bedtime was between 6 and 11pm, it ranged from 6:30pm to 11:30pm during the lockdown. Infants used to be awakened between 0 and 45 minutes during the night before the lockdown, which was then comprised between 0 and 60 minutes during the lockdown. Regular sleeping times on 5-6 days a week were reported for 59.11% of preschool children before the lockdown and 51.82% of preschool children after the lockdown. The number of times they fell asleep in less than 20 minutes 5-6 days a week was 46.56% before lockdown and went down to 39.68%. Before the lockdown, for most preschoolers, bedtimes were regular 5-6 days a week 66.40%, which decreased to 52.63% during the lockdown. For 7.69% they woke up during the night once a week before the lockdown and 9.31% during the lockdown.

Next, the change in sleep from before to during the pandemic was computed. This revealed that more infants experienced a decrease in sleep duration (21.05% increase, 30.89% decrease), a prolongation of sleep latency (34.79% longer, 11.52% shorter), later bedtimes (51.14% later, 13.01% earlier), and more night awakenings (19.86% increase, 13.47% decrease). For preschool children, the shift was in the same direction with less regular sleep duration (6.73% more regular, 17.17% less regular), a more frequent long sleep latency (27.95% more frequent, 8.08% less frequent), more irregular bedtimes (33.67% less regular, 6.39% more regular), and more frequent night awakenings (16.16% more frequent, 8.75% less frequent, Table 2).

**Table 2.** Sleep variables (mean ± standard deviation) in the infant and preschool-age group, and their difference computed as during-before the lockdown from BISQ (infants, reported in minutes) or CSHQ (preschoolers, reported as frequency).

|  | Infants |  |  |  | Preschoolers |  |  |  |
| --- | --- | --- | --- | --- | --- | --- | --- | --- |
|  | Before | During | Change | N | Before | During | Change | N |
| Sleep duration | 770.35 ± 122.07 | 753.69 ± 108.98 | -16.67 ± 97.09 | 296 | 4.26 ± 0.62 | 4.09 ± 0.79 | -0.17 ± 0.67 | 241 |
| Sleep latency | 48.34 ± 127.35 | 58.88 ± 144.72 | 13.31 ± 79.25 | 295 | 3.92 ± 1 | 3.63 ± 1.11 | -0.29 ± 0.85 | 241 |
| Bedtime | 1151.7 ± 248.53 | 1178.98 ± 244.45 | 27.28 ± 258.01 | 297 | 4.12 ± 0.69 | 3.74 ± 0.9 | -0.38 ± 0.84 | 241 |
| Night awakenings | 28.36 ± 88.15 | 31.11 ± 82.35 | 2.75 ± 40.95 | 296 | 2.15 ± 1.27 | 2.25 ± 1.51 | 0.1 ± 1.02 | 241 |

### 3.3. Eating behavior

The eating behavior overall remained similar for the majority, ranging from 56.97% to 73.88% across all categories in infants and preschool children (Table 3). In the remainder, meal size increased in 14.89-17% and decreased in 11.34-16.18% of infants and preschool children.

**Table 3.** Count and percentage for eating behavior variables in infants and preschool children.

| Eating variables | Infants |  |  |  | Preschoolers |  |  |  |
| --- | --- | --- | --- | --- | --- | --- | --- | --- |
|  | Decreased | Similar | Increased | N | Decreased | Similar | Increased | N |
| Meal size | 50 (16.18%) | 213 (68.93%) | 46 (14.89%) | 297 | 28 (11.34%) | 177 (71.66%) | 42 (17%) | 241 |
| Salty snacks | 47 (17.54%) | 198 (73.88%) | 23 (8.58%) | 272 | 47 (19.42%) | 170 (70.25%) | 25 (10.33%) | 241 |
| Sweet snacks | 72 (26.87%) | 180 (67.16%) | 16 (5.97%) | 272 | 91 (37.3%) | 139 (56.97%) | 14 (5.74%) | 241 |
| Fruits | 70 (25.27%) | 168 (60.65%) | 39 (14.08%) | 263 | 48 (19.43%) | 161 (65.18%) | 38 (15.38%) | 236 |
| Vegetables | 34 (12.27%) | 204 (73.65%) | 39 (14.08%) | 263 | 32 (12.96%) | 180 (72.87%) | 35 (14.17%) | 238 |

The consumption of Salty snacks increased in 8.58-10.33% and decreased in 17.54-19.42% of children.

For Sweet snacks, an increase was observed in 5.74-5.97% and a decrease in 26.87-37.3% for both age groups. Children increased their consumption of Fruits in 14.08-15.38% of cases, while 19.43-25.27% decreased it. Vegetables eating increased in 14.08-14.17% of children and decreased in 12.27-12.96%.

### 2.4. Social isolation and eating behavior

We then examined whether the extent of social contact related to eating behavior in both age groups. Overall, neither the change in meal size, nor snacks, fruits and vegetables were related to the infants’ extent of social contacts (Table 4, all p>0.05). In preschool children, a trend was observed in the association between change in meal size and social contact through quarantine status (p=0.07). In other words, this would indicate that preschool children in quarantine were more likely to increase their meal size, compared to non-quarantined preschool children (Figure 1). The other measures of social contact were neither related to meal size nor to type of consumed food (Table 5, all p>0.05).

**Table 4.** Association between extent of social contact and eating behavior in infants. Unstandardized beta coefficients (b) and corrected p-values (p) are indicated from the generalized linear model. Significant associations (p < 0.05) are presented in bold.

|  | Infants |  |  |  |  |  |  |  |  |  |
| --- | --- | --- | --- | --- | --- | --- | --- | --- | --- | --- |
|  | Meal size |  | Vegetables |  | Fruits |  | Salty snacks |  | Sweet snacks |  |
|  | b | p | b | p | b | p | b | p | b | p |
| Household size | 0.057 | 0.195 | -0.028 | 0.831 | -0.080 | 0.331 | 0.019 | 0.833 | -0.059 | 0.543 |
| Quarantine status | 0.015 | 0.828 | -0.008 | 0.919 | -0.090 | 0.525 | 0.023 | 0.833 | 0.072 | 0.543 |
| % Social activities | 0.133 | 0.484 | -0.404 | 0.143 | -0.218 | 0.525 | 0.251 | 0.559 | 0.161 | 0.543 |
| Sex | 0.054 | 0.484 | -0.136 | 0.155 | -0.135 | 0.331 | 0.115 | 0.536 | -0.029 | 0.728 |
| Age | 0.217 | **<0.001** | 0.214 | **<0.001** | 0.144 | 0.192 | 0.020 | 0.833 | -0.143 | 0.113 |
| Parental stress | 0.025 | 0.484 | -0.014 | 0.857 | -0.015 | 0.721 | 0.008 | 0.833 | 0.033 | 0.543 |

**Table 5.**
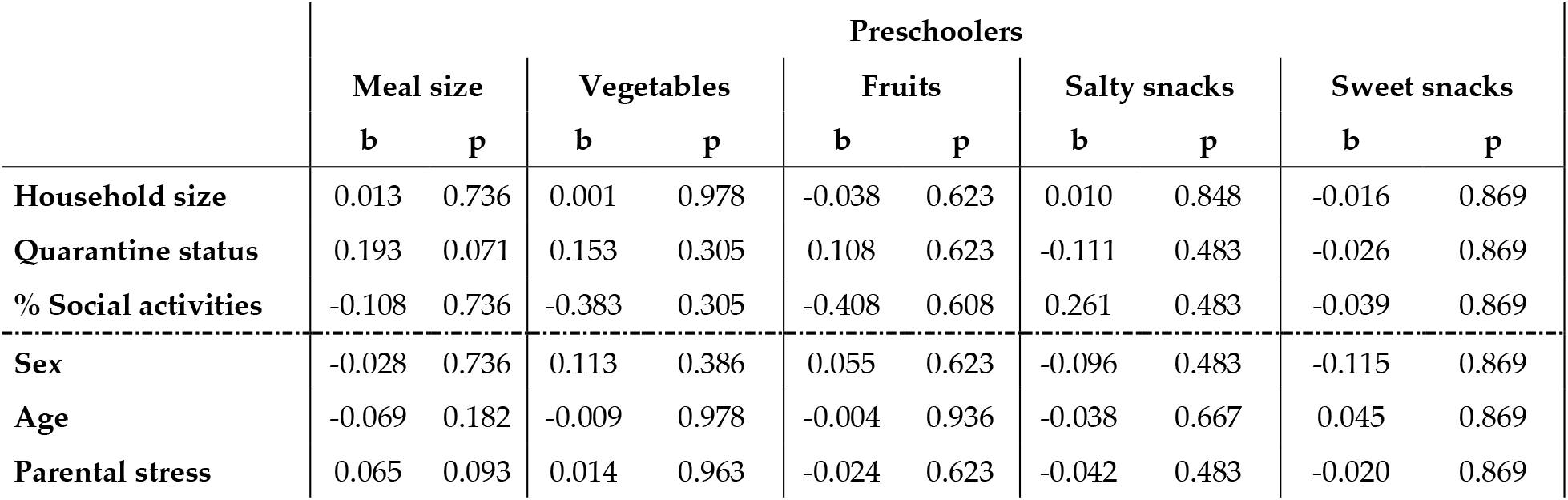
Association between extent of social contact and eating behavior in preschool children. Unstandardized beta coefficients (b) and corrected p-values (p) from the generalized linear model.

|  | Preschoolers |  |  |  |  |  |  |  |  |  |
| --- | --- | --- | --- | --- | --- | --- | --- | --- | --- | --- |
|  | Meal size |  | Vegetables |  | Fruits |  | Salty snacks |  | Sweet snacks |  |
|  | b | p | b | p | b | p | b | p | b | p |
| Household size | 0.013 | 0.736 | 0.001 | 0.978 | -0.038 | 0.623 | 0.010 | 0.848 | -0.016 | 0.869 |
| Quarantine status | 0.193 | 0.071 | 0.153 | 0.305 | 0.108 | 0.623 | -0.111 | 0.483 | -0.026 | 0.869 |
| % Social activities | -0.108 | 0.736 | -0.383 | 0.305 | -0.408 | 0.608 | 0.261 | 0.483 | -0.039 | 0.869 |
| Sex | -0.028 | 0.736 | 0.113 | 0.386 | 0.055 | 0.623 | -0.096 | 0.483 | -0.115 | 0.869 |
| Age | -0.069 | 0.182 | -0.009 | 0.978 | -0.004 | 0.936 | -0.038 | 0.667 | 0.045 | 0.869 |
| Parental stress | 0.065 | 0.093 | 0.014 | 0.963 | -0.024 | 0.623 | -0.042 | 0.483 | -0.020 | 0.869 |

**Figure 1.**
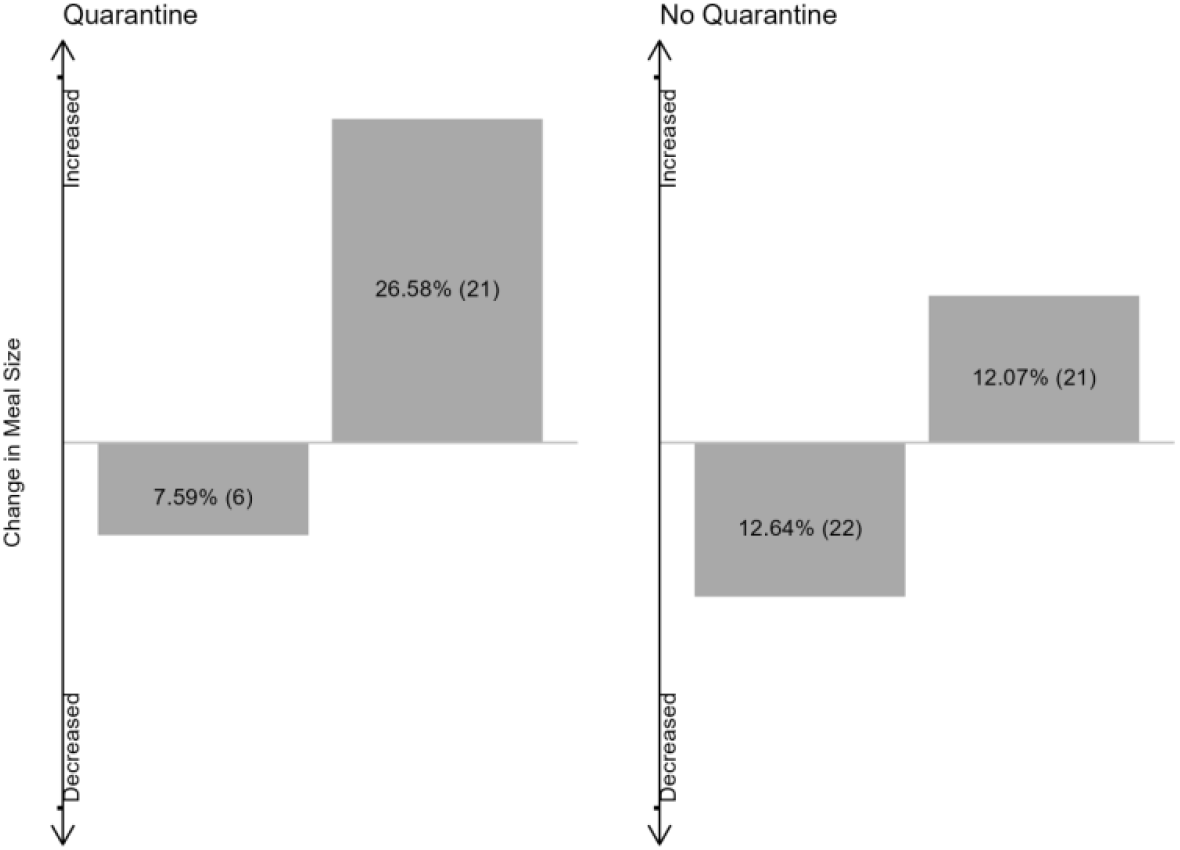
Change in meal size for the groups of quarantined and non-quarantined preschool children. Numbers refer to percentages and counts of preschool children for whom a change in meal size was reported.

The control variables included in our model (age, sex and parental stress) were not related to the children’s eating behavior, with the exception that older children decreased meal size to a larger extent from before to during the pandemic (b = -0.217, p < 0.001), and ate more vegetables (b = 0.214, p < 0.001).

### 3.5. Social isolation and sleep behavior

We then examined whether the extent of social contact was related to children’s sleep behavior. Neither in infants nor in preschool children was the extent of social contact associated with sleep variables (i.e., duration, sleep latency, bedtimes nor with the number of nighttime awakenings; Table 6, all p>0.05). Change in sleep behavior was not related to age, sex and parental stress, except for a link between parental stress and sleep latency in preschool children, such that increased stress related to longer sleep latency (b = 0.130, p = 0.043).

**Table 6.** Extent of social contact association with sleep behavior. Unstandardized beta coefficients (b) and corrected p-values (p) from the linear mixed model.

|  | Infants |  |  |  |  |  |  |  |
| --- | --- | --- | --- | --- | --- | --- | --- | --- |
|  | Sleep duration |  | Sleep latency |  | Bedtimes |  | Nighttime awakenings |  |
|  | b | p | b | p | b | p | b | p |
| Household size | 5.846 | 0.334 | -6.137 | 0.521 | 7.993 | 0.760 | -1.705 | 0.895 |
| Quarantine status | -20.355 | 0.107 | 19.213 | 0.222 | -31.804 | 0.760 | 10.324 | 0.174 |
| % Social activities | -51.058 | 0.107 | -2.123 | 0.932 | 38.103 | 0.760 | 1.655 | 0.895 |
| Sex | -4.342 | 0.745 | 1.088 | 0.932 | -13.249 | 0.760 | 1.184 | 0.895 |
| Age | 13.975 | 0.107 | -2.401 | 0.932 | -48.104 | 0.088 | 7.735 | 0.095 |
| Parental stress | 5.541 | 0.334 | -9.111 | 0.222 | -1.543 | 0.908 | -0.773 | 0.895 |
|  | Preschoolers |  |  |  |  |  |  |  |
|  | Sleep duration |  | Sleep latency |  | Bedtimes |  | Nighttime awakenings |  |
|  | b | p | b | p | b | p | b | p |
| Household size | 0.043 | 0.746 | 0.029 | 0.722 | 0.065 | 0.622 | 0.007 | 0.933 |
| Quarantine status | -0.043 | 0.746 | -0.249 | 0.139 | 0.271 | 0.117 | -0.049 | 0.933 |
| % Social activities | 0.234 | 0.746 | -0.221 | 0.636 | -0.222 | 0.622 | 0.491 | 0.848 |
| Sex | 0.042 | 0.746 | 0.039 | 0.722 | -0.096 | 0.622 | -0.023 | 0.933 |
| Age | 0.001 | 0.978 | -0.058 | 0.636 | -0.003 | 0.965 | -0.076 | 0.848 |
| Parental stress | 0.067 | 0.453 | 0.130 | 0.043 | -0.099 | 0.117 | -0.057 | 0.848 |

Overall, the results indicate only a negligible relationship between the extent of social contact with eating behavior in infant and preschool children during the COVID-19 pandemic. This was indicated by a trend suggesting that being in quarantine related to increased meal size in preschoolers.

## 4. Discussion

Social isolation in adults can be related to poor sleep (Azizi-Zeinalhajlou et al. 2022; Cacioppo et al. 2002), and altered eating behavior (Mason et al. 2016; Wright and Pritchard 2009). Yet, the interplay between those three factors remains to be elucidated in early childhood, which was evaluated with this observational study during the COVID-19 confinement. Specifically, parents of 309 infants and 253 preschool children provided information before and during the confinement, rating their children’s social contacts, eating behavior and sleep. Overall, our results indicate that children’s social contact is not strongly associated with sleeping behavior, nor with eating behavior, with the exception for a trend such that quarantined preschool children tended to have increased meal size compared to non-quarantined ones. The nuanced findings emphasize the need to better understand which aspect of social isolation is impactful on children’s health through changes in their sleeping and eating behavior.

Contrary to our hypotheses, meal size, as well as the consumption of snacks, fruits and vegetables, was not related to the extent of social contact, suggesting that factors beyond social isolation play a role in dynamics of eating behavior in infants and preschoolers. We observed that preschoolers being in quarantine had a heightened chance of increasing meal size in relation to the confinement, yet this association remained as a trend after adjusting for multiple comparisons. This relationship is concordant with findings demonstrating increased subjective isolation in obese adolescents (Mason, 2020). Interestingly, our observation prevailed only in preschool children, yet not in infants, which could point to age-specific differences in need for social contact. Increased meal size and more unhealthy eating behaviors during the pandemic have been reported in children 6 years and older, suggesting that the effect of social isolation may be dependent on age (González-Monroy et al. 2021; Pujia et al. 2021). Age effects could also be attributed to the extent of contacts with people outside of the immediate household. For infants, the main social contacts are their primary caregiver(s), while preschool children are more likely to have extended contacts with other adults. Thus, as the change in social contact with primary caregiver(s) during the pandemic did not change, infants’ changes were probably less extreme. Further, we lacked information on other aspects of eating behavior such as the timing of meals which could also be affected by the pandemic and is related to sleeping behavior (Mühlematter et al. 2023). Moreover, eating behavior is the primary factor determining the composition of the gut microbiota, and interestingly animal research observed associations between social isolation, the gut microbiota and the brain (Dunphy-Doherty et al. 2018; Lopizzo et al. 2021). Thus, the gut microbiota could be an interesting candidate in future research on the interplay between social isolation, sleeping and eating behavior. Our results show that only quarantine could have an effect on eating behavior, and only in children aged 3 to 6 years old.

In line with previous studies (Markovic et al., 2021), sleep behavior shifted towards a decreased sleep quality (i.e., shorter sleep duration, longer sleep latency, more nighttime awakenings). Similar to eating, children’s changes in sleep variables were not strongly linked to the degree of their social contacts. Interestingly, this contrast with findings in adults. Effects of adults’ social isolation on sleep quality are well-documented, showing, for example that subjective loneliness often accompanies subjective poor sleep quality (Bhatti and Haq 2017; Cacioppo et al. 2002; McLay et al. 2021). Thus, the difference to children’s sleep might be attributable to other factors, such as parents engagement in mindfulness techniques or the presence of pets, as previously demonstrated (Markovic et al. 2021). Another possibility is that children’s interactions with their parents sufficiently safeguarded them from potential negative effects of reduced social contact on their sleep. Moreover, our study used objective ratings of social contact, whereas studies involving adults relied on individual perceptions of isolation (Cacioppo et al. 2002; Cacioppo, Hawkley, and Thisted 2010). Thus, the adverse outcomes of social isolation might be subjectively biased in adults, making results not directly comparable. To summarize our findings, sleep behavior change during the pandemic was not dependent on externally reported measures of social contact.

Age, sex, and parental stress had a small impact on children’s both eating behavior and sleep in our dataset. Higher parental stress marginally increased the chance of increased meal size in the preschool group. Thus, parental stress, depending on the age group, remains an important factor to consider in the framework of children’s eating behavior. In alignment with this, stress exposure in adults can increase food intake, with specifically increasing the preference for high-fat, high-sugar foods (Tomiyama 2019). Future investigations to elucidate the impact of stress on young children’s eating behaviors could inform strategies for mitigation.

In conclusion, this study sheds light on the association of social isolation with sleeping and eating behavior in children up to 6 years old. We report that, in contrast to findings in adolescents and adults, social contact beyond the family household is only negligibly associated with young children’s sleep and eating behavior. Thus, more research is needed to gain a deeper understanding of the maturational transition and the relationships between social contact, sleep patterns and eating behavior. This is a significant concern, particularly in light of the rising rates of obesity (Pan, Wang, and Pan 2021) and sleep problems in children observed in recent decades (Chen et al. 2021). While these issues have complex causes, social isolation might potentially contribute to them, making it a worthwhile focus for addressing mental, metabolic and chronobiological health.

## Author Contributions

CM, SK, AM, and MB designed and conducted the study. CM performed the analyses. CM and SK wrote the manuscript. All authors discussed the results and contributed to the final manuscript.

## Data Availability Statement

Ethical restrictions (Institutional ethics board, University of Fribourg) apply to this paper, which prevents the public sharing of individual data that contain potentially sensitive information.

## Acknowledgments

We thank Océane Minot, Debora Castiglioni, Valentine Horii, and Dr. Maria Berrozpe for assistance in data collection. Our appreciation goes to the participants who contributed to the study.

## Appendix A Activity Times Questionnaire

Since the lockdown, what kind of activities does your child engage in? We are interested in the things you do together with the child (or the child does with siblings or friends) referred to as “together” and the activities the child does “alone”. Please indicate the number of hours (hours:minutes) on an average day.

**Watching television** Together:___ Alone:___

**Playing game on TV/tablet** Together:___ Alone:___

**Using pedagogic apps** Together:___ Alone:___

**Doing manual activities (e.g. colouring, cooking)** Together:___ Alone:___

**Read something** Together:___ Alone:___

**Calling someone** Together:___ Alone:___

**Board game** Together:___ Alone:___

**Playing with toy(s)** Together:___ Alone:___

**Playing outside** Together:___ Alone:___

**Chores and household tasks** Together:___ Alone:___

**Other**: Together:___ Alone:___

## Disclaimer/Publisher’s Note

The statements, opinions and data contained in all publications are solely those of the individual author(s) and contributor(s) and not of MDPI and/or the editor(s). MDPI and/or the editor(s) disclaim responsibility for any injury to people or property resulting from any ideas, methods, instructions or products referred to in the content.

